# A Novel Abnormality Annotation Database for COVID-19 Affected Frontal Lung X-rays

**DOI:** 10.1101/2021.01.07.21249323

**Authors:** Surbhi Mittal, Vasantha Kumar Venugopal, Vikash Kumar Agarwal, Manu Malhotra, Jagneet Singh Chatha, Savinay Kapur, Ankur Gupta, Vikas Batra, Puspita Majumdar, Aakarsh Malhotra, Kartik Thakral, Saheb Chhabra, Mayank Vatsa, Richa Singh, Santanu Chaudhury

## Abstract

**Purpose:** To advance the usage of CXRs as a viable solution for efficient COVID-19 diagnostics by providing large-scale annotations of the abnormalities in frontal CXRs in BIMCV-COVID19+ database, and to provide a robust evaluation mechanism to facilitate its usage.

**Materials and Methods:** We provide the abnormality annotations in frontal CXRs by creating bounding boxes. The frontal CXRs are a part of the existing BIMCV-COVID19+ database. We also define four different protocols for robust evaluation of semantic segmentation and classification algorithms. Finally, we benchmark the defined protocols and report the results using popular deep learning models as a part of this study.

**Results:** For semantic segmentation, Mask-RCNN performs the best among all the models with a DICE score of 0.43 ± 0.01. For classification, we observe that MobileNetv2 yields the best results for 2-class and 3-class classification. We also observe that deep models report a lower performance for classifying other classes apart from the COVID class.

**Conclusion:** By making the annotated data and protocols available to the scientific community, we aim to advance the usage of CXRs as a viable solution for efficient COVID-19 diagnostics. This large-scale data will be useful for ML algorithms and can be used for learning radiological patterns observed in COVID-19 patients. Further, the protocols will facilitate ML practitioners for unified large-scale evaluation of their algorithms.

**Data Availability Statement:** The data associated with this work is available here : <u>Radiologists’ Annotations on COVID-19+ X-rays https://osf.io/b35xu/ via @OSFramework and</u> http://covbase4all.igib.res.in/.

## Introduction

In the face of the Coronavirus pandemic, more than 65 million people have been exposed to the COVID-19 virus, globally(1). To contain the virus’s spread, it is necessary to execute contact tracing, mass screening, and testing of the suspected patients. Radiological investigations, especially X-ray and CT scans, can play a major role in the diagnostic workflow of patients(2,3). COVID-19 pneumonia has a characteristic spectrum of findings on chest x-rays which are bilateral peripheral patchy opacities in predominant lower lobe distributions(4,5). While these findings may overlap with viral or other atypical pneumonia(6,7), they are distinctively different from the more common typical pneumonia of bacterial origin. These differences offer an opportunity to use x-rays for screening and diagnosis. Some of the differences can be observed in Figure 1. For differentiating COVID-19 pneumonia, frontal CXRs remain a popular choice for different studies(8–11). These studies utilize existing databases of COVID-19 affected CXRs(11-13). However, these datasets are small and lack abnormality localization information. The evaluation of COVID-19 pneumonia classifiers also suffers from a lack of well-defined validation frameworks and unavailability of common testing sets. Moreover, different algorithms have divergent problem statements classifying in different numbers and types of classes (COVID, non-COVID pneumonia, normal, etc).

**Figure 1.**
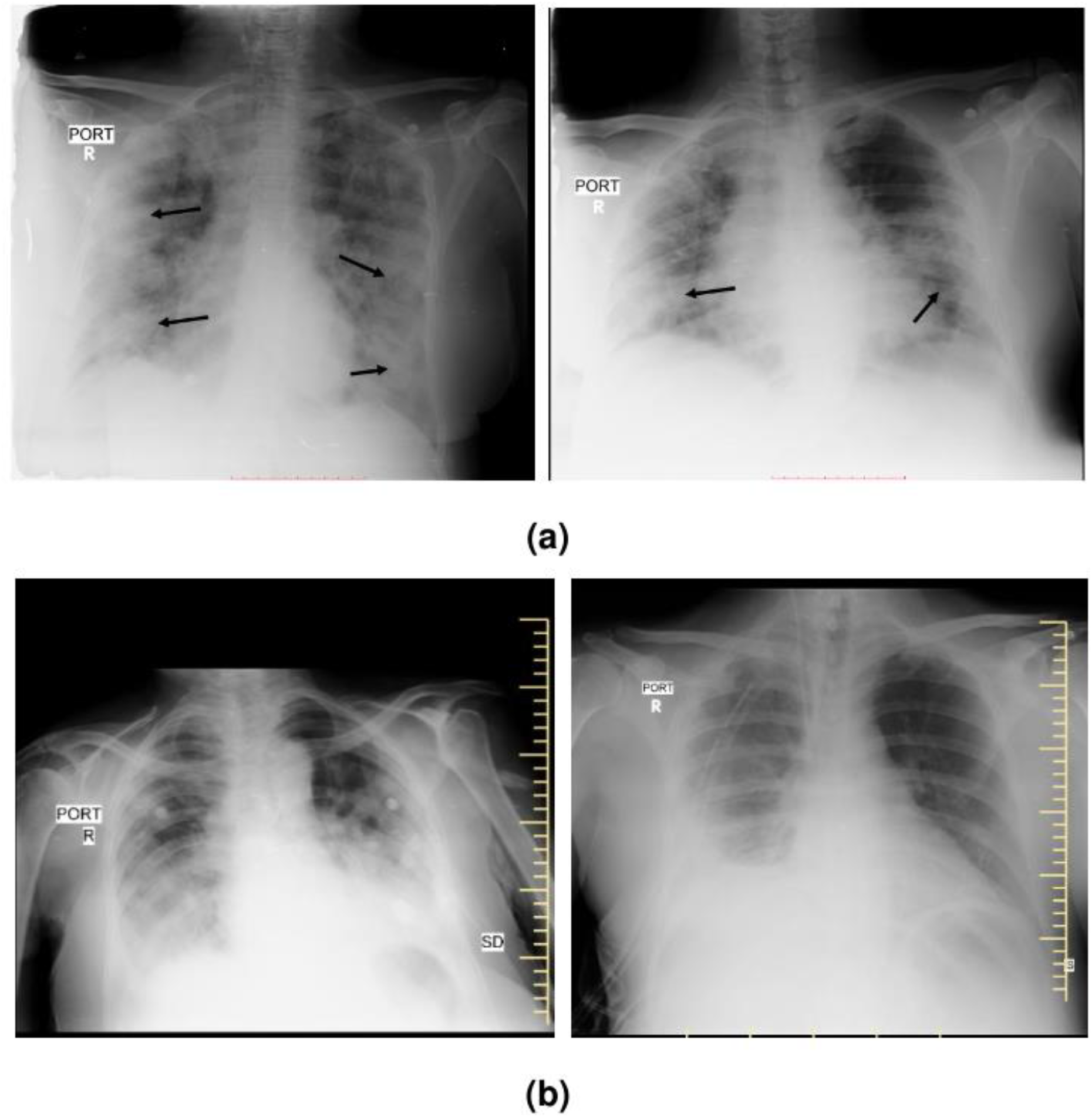
Chest x-rays of (a) RT-PCR proven COVID-19 pneumonia in cases showing the typical bilateral peripheral consolidations and ground-glass opacities. (b) Non-COVID-19 pneumonia in cases showing the lobar distribution of consolidation with pleural effusion.

In this study, we introduce the COVID Abnormality Annotation for X-Rays (CAAXR) database. The proposed CAAXR database builds upon the BIMCV-COVID19+ database(12) by providing abnormality localization of frontal CXRs. The marked annotations include a comprehensive set of abnormalities, including but not limited to atelectasis, consolidation, pleural effusion, and edema. On 1749 different x-rays from the BIMCV-COVID19+ database, we mark a total of 3943 abnormalities as bounding boxes. Further, our study creates a robust evaluation mechanism to facilitate the usage of the BIMCV-COVID19+ database with the proposed annotations. We design protocols that provide a four-way testing mechanism on a consolidated set of x-rays. These protocols include one protocol for semantic segmentation, and a protocol each for two-, three-, and four-class classification. These protocols also incorporate cross-validation, allowing researchers to check the stability of their algorithms. Lastly, we establish the baseline performance for popular deep learning algorithms with the specified protocols.

## Materials and Methods

### Data Sources

There are two data sources- BIMCV-COVID19+ dataset(12) and CheXpert dataset(15). BIMCV-COVID19+ dataset is a large dataset of CXRs and CT images of COVID-19 positive patients. It contains 2265 CXR images. The dataset contains multiple studies of the same subject and multiple images per study. Additionally, the dataset contains phenotypic data and metadata information for acquired images. This study focuses only on frontal CXRs from the dataset. More details about the dataset can be found in the preprint released with it(14). The CheXpert dataset is a large chest x-ray dataset containing 223,414 images. It is a popular dataset for performing automatic CXR interpretation, containing radiologist-labeled data. Based on the radiological findings, each x-ray image is labeled either 0(negative), 1(positive), or ‘u’(uncertain) for 14 pre-defined classes. For this study, a fixed subset of 7,212 images are acquired from it. Images in the BIMCV-COVID19+ dataset are 12-bit grayscale high-resolution PNG images. The images are pre-processed by converting the PNG files into standard 8-bit image format. The metadata information specifies images that need to be contrast-inverted for proper viewing. The key “20500020” is checked in the corresponding JSON metadata file for each image. If the key exists with the value “INVERSE”, contrast-inversion is performed on the corresponding CXR image. All images are resized to a fixed resolution of 224×224 using bi-cubic interpolation. For the CheXpert dataset, the downsampled resolution CheXpert-v1.0 dataset is used. It has images of resolution 300×300 pixels in the PNG format. The images are pre-processed by resizing to a fixed resolution of 224×224 using bicubic interpolation.

### Ground Truth

A total of 1,749 chest X-Rays have been annotated by six radiologists each with more than years’ experience of reading chest X-rays. The annotations are created in the form of rectangular bounding boxes and have associated finding labels including Consolidation, Atelectasis, Pleural Effusion among others. Non-lung abnormalities like fracture, cardiomegaly, and presence of support devices are also noted. The images are downloaded from the BIMCV-COVID19+ dataset(12) in PNG format, converted to DICOM, and associated metadata of each image (available as a corresponding JSON) loaded into the DICOM file. The subsequently produced DICOM files are loaded onto the CARPL Annotation Platform (CARING, Delhi, INDIA) and accessed by radiologists who drew bounding boxes around each image. At the study level, the images are annotated for ‘Normal/Abnormal’ classification and for image quality on a scale of ‘1-3’ (1- poor, 2-acceptable, and 3-good quality). Some samples for annotations can be seen in Figure 2.

**Figure 2.**
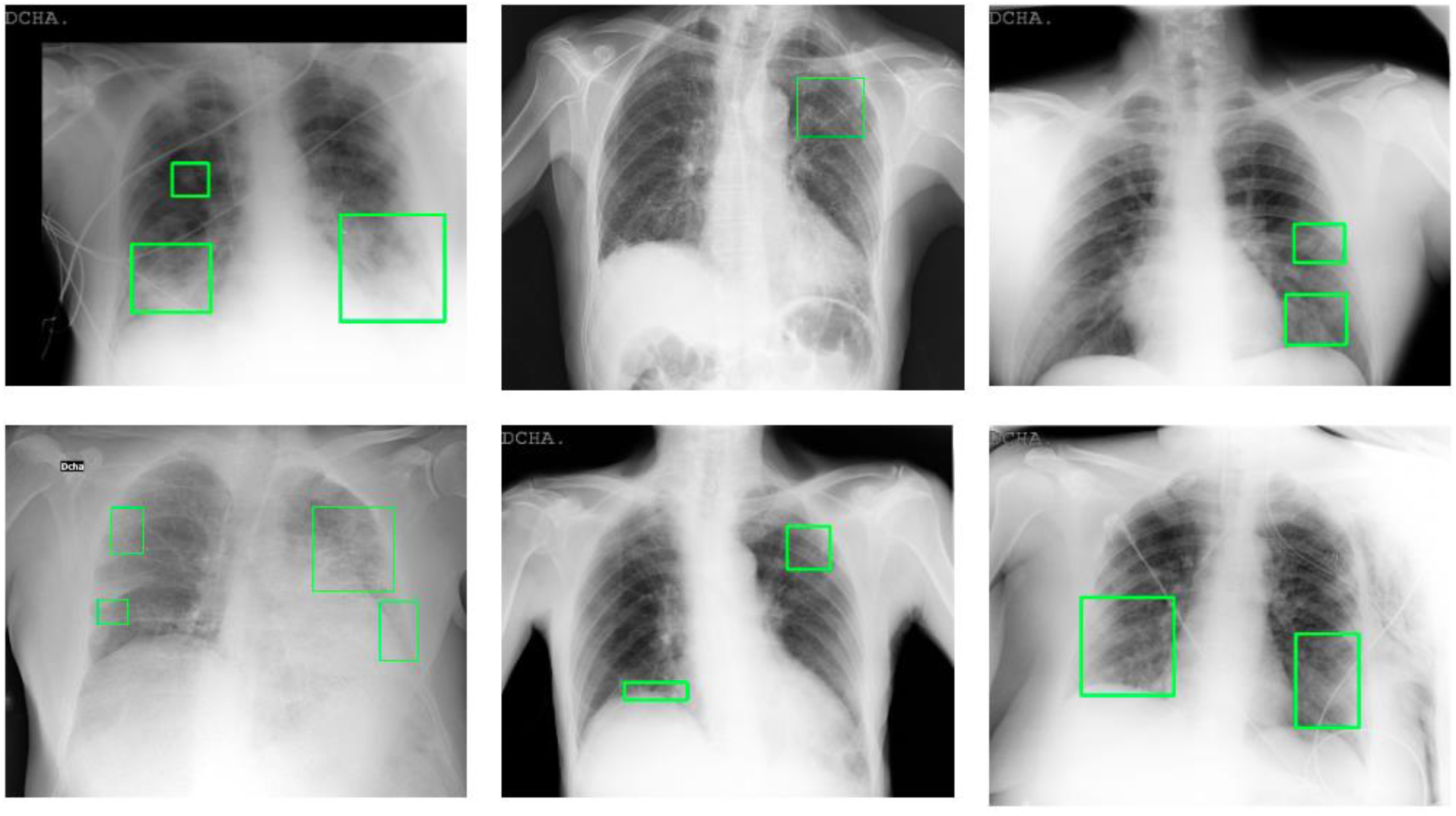
Samples of abnormality annotations performed for BIMCV-COVID19+ dataset.

### Experimental Design

As a part of this study, we introduce four protocols for evaluation. A total of 3,288 samples are drawn from the BIMCV-COVID19+, and a total of 7,212 samples are drawn from the CheXpert across all protocols. All protocols use 2-fold cross-validation. The data is split into an approximate 40-10-50 train, validation, and test split. The 50% data used for testing is consistent across the cross-validation experiments and the remaining 50% is divided into a train and validation set. Data related to the protocols has been summarized in Figure 3.

**Figure 3.**
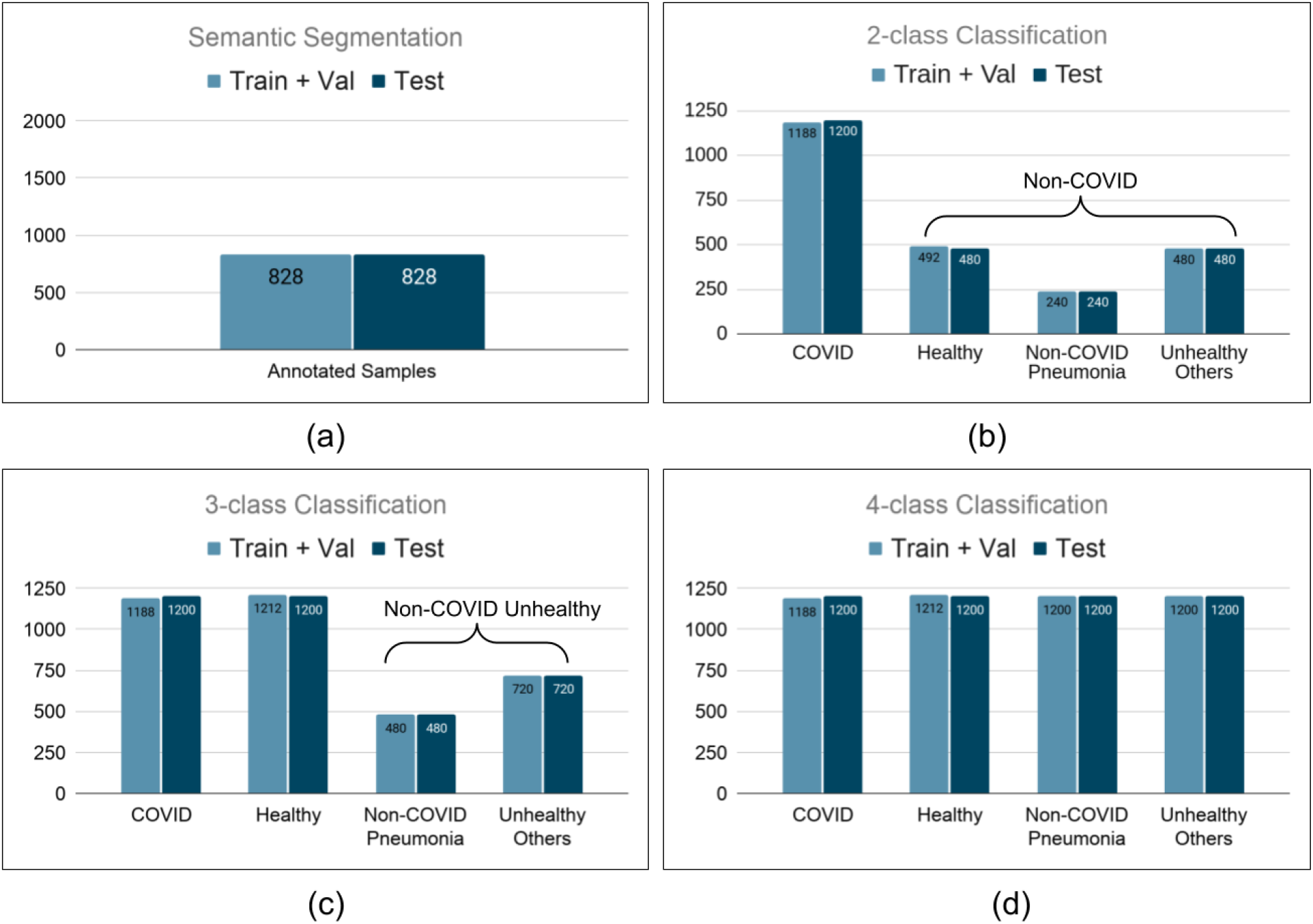
Number of samples present in protocol for (a) Semantic Segmentation. (b) 2-class Classification. (c) 3-class Classification. (d) 4-class Classification.

- Semantic segmentation For this protocol, a total of 1,656 samples from the BIMCV-COVID19+ dataset are drawn. The bounding box annotations for the samples are used to mark the unhealthy regions on the x-rays by creating a binary mask using the bounding box coordinates. These masked images are resized to a resolution of 224×224. The samples are randomly split into training and testing.
- 2-class classification (COVID/Non-COVID) In this protocol, images belong to one of two classes- COVID or Non-COVID. 2,388 CXR images for COVID positive subjects are obtained from the BIMCV-COVID19+ dataset, while 2,412 Non-COVID infected CXR images are obtained from the CheXpert dataset. The Non-COVID infected CXRs mages are either healthy, infected with non-COVID pneumonia, or have some other abnormalities. The testing set contains 2400 samples.
- 3-class classification (COVID/Non-COVID Unhealthy/Healthy) In this protocol, x-rays belong to one of the three classes- COVID, Non-COVID Unhealthy, or Healthy. The set of COVID 2,388 CXRs used in the previous protocol is used in this protocol. However, this time a subset of 4,812 x-rays is obtained from the CheXpert dataset for the Non-COVID Unhealthy and Healthy class samples. The samples in the Non-COVID Unhealthy class consist of CXRs with Non-COVID pneumonia or with other radiological abnormalities.
- 4-class classification (COVID/Non-COVID Pneumonia/Unhealthy Other/Healthy) In this protocol, x-rays belong to one of four classes- COVID, Non-COVID Pneumonia, Unhealthy Others, or Healthy. The set of COVID 2,388 chest x-rays used in all classification protocols is the same as in the previous two protocols. A subset of 7,212 images is obtained from the CheXpert dataset for samples belonging to the Non-COVID Pneumonia, Unhealthy Others, and Healthy classes.

### Models and Training

This section provides information about baseline models and their implementation. The hyperparameters for the different models are selected based on observing their performance on the validation sets.

For semantic segmentation, we evaluate three segmentation models-UNet(17), SegNet(18), and Mask-RCNN(19). Training of UNet and SegNet models is performed on Python 3.6.9 on Linux, kernel 5.3.0-61-generic 18.04.1-Ubuntu SMP, architecture x86_64. The models are trained using PyTorch (v.1.4.0) with the input size of images as 224×224. The training batch size is set as 4. The model is trained for 25 epochs using Adam optimizer with an initial learning rate of 0.0001. The Binary Cross-Entropy loss is minimized. A system with Nvidia RTX 2080Ti and IntelXeon (having 48 cores) with 128 GB RAM is used for training the above models. For Mask-RCNN, implementation is done in PyTorch (1.6.0+cu101) with input images of the same size. The training batch size is set to 16. The model is trained for 25 epochs using SGD optimizer with learning rate set to 0.001 and momentum to 0.9. A combination of classification, localization, and segmentation mask loss are minimized where Binary Cross-Entropy loss is used for classification and segmentation task, and Smooth L1 loss for the classification task. The training is performed on Google Colab with Nvidia Tesla T4 as the GPU accelerator. In order to evaluate the classification performance, four deep learning models namely-DenseNet121(20), MobileNetv2(21), ResNet18(22), and VGG19(23) are trained separately using two-fold cross-validation for each of the classification protocols. The classification models are trained by adding two fully connected dense layers of 1024 and 512 dimensions after the final convolutional layer of each model. During the experiments, convolutional layers of the models are frozen and only the dense layers are updated using cross-entropy loss function. Models are trained for 30 epochs with Stochastic Gradient Descent (SGD) optimizer. The learning rate is set to 0.001 and momentum to 0.9. A batch size of 10 is used during training. The code is implemented in PyTorch (v1.4.0). All the experiments are performed on a system with Nvidia RTX 2080Ti GPU and IntelXeon (having 48 cores) with 128 GB RAM.

### Evaluation

The performance for Semantic Segmentation is evaluated using two metrics-Intersection over Union (IoU) and DICE coefficient. The mean value of the respective metrics is calculated by averaging their values over the entire dataset. The results for classification are evaluated using class-wise AUC for 2-class, 3-class, and 4-class classification for the deep models. Additionally, the average performance of the models is evaluated in terms of sensitivity at 99% and 90% specificity. With new algorithms focusing on the quantification and tracking progression of COVID-19 pneumonia, we also evaluate the models on a subset of the test set. This subset contains 768 x-rays with bounding box annotations for consolidation and ground-glass opacities only and provides a benchmark for pneumonia and other regions’ segmentation within the context of COVID-19.

## Results

All experiments are performed using 2-fold cross-validation. The results for segmentation on the test set, and the consolidation test subset are shown in Table 1. We observe that Mask-RCNN performs the best among all the models with an IoU of 0.30 ± 0.01 and a DICE score of 0.43 ± 0.01 on the test set. Results for a few instances are shown in Figure 4. The results of classification are summarized in Table 2. We observe that MobileNetv2 yields the best results for 2-class and 3-class classification. For 3-class and 4-class classification, the performance of the models for classifying other classes apart from the COVID class is low. This could be due to the small number of samples present in the dataset to learn large diversity present in these classes. Table 2(a) shows the AUC for 2-class, 3-class, and 4-class classification using the four deep models. It is observed that all the models achieve high performance for classifying the COVID class corresponding to all the protocols. This high performance could be due to difference in dataset properties as COVID and non-COVID samples are taken from two different datasets. This might include learning source properties of the x-ray machines, or, digital signatures introduced due to software-level image manipulations. This observation is strengthened by the fact the models have a subpar performance for classification within the classes from the Chexpert dataset, while the COVID-19 classification is consistently very good.

**Table 1.**
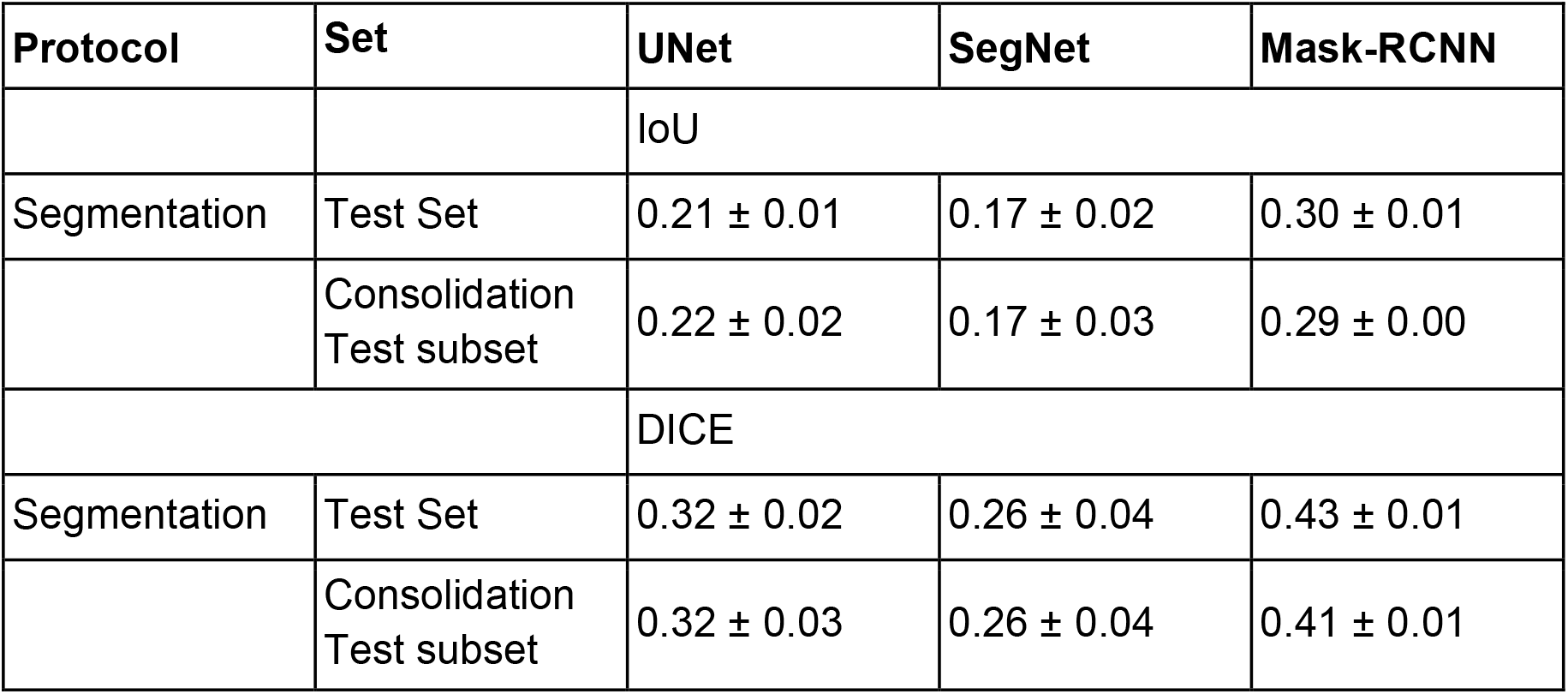
Evaluation of existing deep learning models for COVID-19 semantic segmentation.

| Protocol | Set | UNet | SegNet | Mask-RCNN |
| --- | --- | --- | --- | --- |
|  |  | IoU |  |  |
| Segmentation | Test Set | $0.21 \pm 0.01$ | $0.17 \pm 0.02$ | $0.30 \pm 0.01$ |
| | Consolidation<br>Test subset | $0.22 \pm 0.02$ | $0.17 \pm 0.03$ | $0.29 \pm 0.00$ |
|  |  | DICE |  |  |
| Segmentation | Test Set | $0.32 \pm 0.02$ | $0.26 \pm 0.04$ | $0.43 \pm 0.01$ |
| | Consolidation<br>Test subset | $0.32 \pm 0.03$ | $0.26 \pm 0.04$ | $0.41 \pm 0.01$ |

**Table 2.** (a) Class-wise AUC corresponding to existing deep learning algorithms for all classification protocols. (b) Evaluation of existing deep learning algorithms for COVID-19 prediction corresponding to all classification protocols. In the case of multi-class classification, a one-vs-all strategy is used to report sensitivity for COVID detection.

| Protocol | Classes | DenseNet121 | MobileNetv2 | ResNet18 | VGG19 |
| --- | --- | --- | --- | --- | --- |
|  |  | AUC |  |  |  |
| 2-class classification | COVID | 0.997 ± 0.000 | 0.998 ± 0.000 | 0.997 ± 0.000 | 0.996 ± 0.001 |
| | Non-COVID | 0.998 ± 0.002 | 0.999 ± 0.000 | 0.998 ± 0.000 | 0.997 ± 0.001 |
| 3-class classification | COVID | 0.997 ± 0.000 | 0.998 ± 0.000 | 0.997 ± 0.000 | 0.996 ± 0.000 |
| | Non-COVID Unhealthy | 0.895 ± 0.004 | 0.897 ± 0.001 | 0.889 ± 0.000 | 0.892 ± 0.001 |
| | Healthy | 0.895 ± 0.006 | 0.900 ± 0.001 | 0.889 ± 0.001 | 0.889 ± 0.001 |
| 4-class classification | COVID | 0.997 ± 0.000 | 0.997 ± 0.000 | 0.997 ± 0.000 | 0.995 ± 0.000 |
| | Non-COVID Pneumonia | 0.788 ± 0.004 | 0.790 ± 0.006 | 0.757 ± 0.015 | 0.785 ± 0.002 |
| | Unhealthy Others | 0.785 ± 0.003 | 0.794 ± 0.004 | 0.774 ± 0.007 | 0.797 ± 0.002 |
| | Healthy | 0.862 ± 0.001 | 0.869 ± 0.002 | 0.844 ± 0.007 | 0.852 ± 0.002 |
| Protocol |  | DenseNet121 | MobileNetv2 | ResNet18 | VGG19 |
|  |  | Sensitivity @ 99% Specificity |  |  |  |
| 2-class classification | | 95.16 ± 0.35 | 98.91 ± 0.11 | 96.62 ± 0.29 | 94.04 ± 0.76 |
| 3-class classification | | 95.50 ± 0.17 | 97.95 ± 0.47 | 96.27 ± 0.14 | 94.06 ± 0.08 |
| 4-class classification | | 94.75 ± 0.74 | 97.43 ± 0.33 | 96.69 ± 1.37 | 92.47 ± 0.51 |
|  |  | Sensitivity @ 90% Specificity |  |  |  |
| 2-class classification | | 99.87 ± 0.05 | 99.95 ± 0.50 | 99.79 ± 0.05 | 99.50 ± 0.23 |
| 3-class classification | | 99.89 ± 0.02 | 99.89 ± 0.02 | 99.79 ± 0.00 | 99.50 ± 0.11 |
| 4-class classification | | 99.91 ± 0.03 | 99.81 ± 0.01 | 99.68 ± 0.01 | 99.33 ± 0.11 |

**Figure 4.**
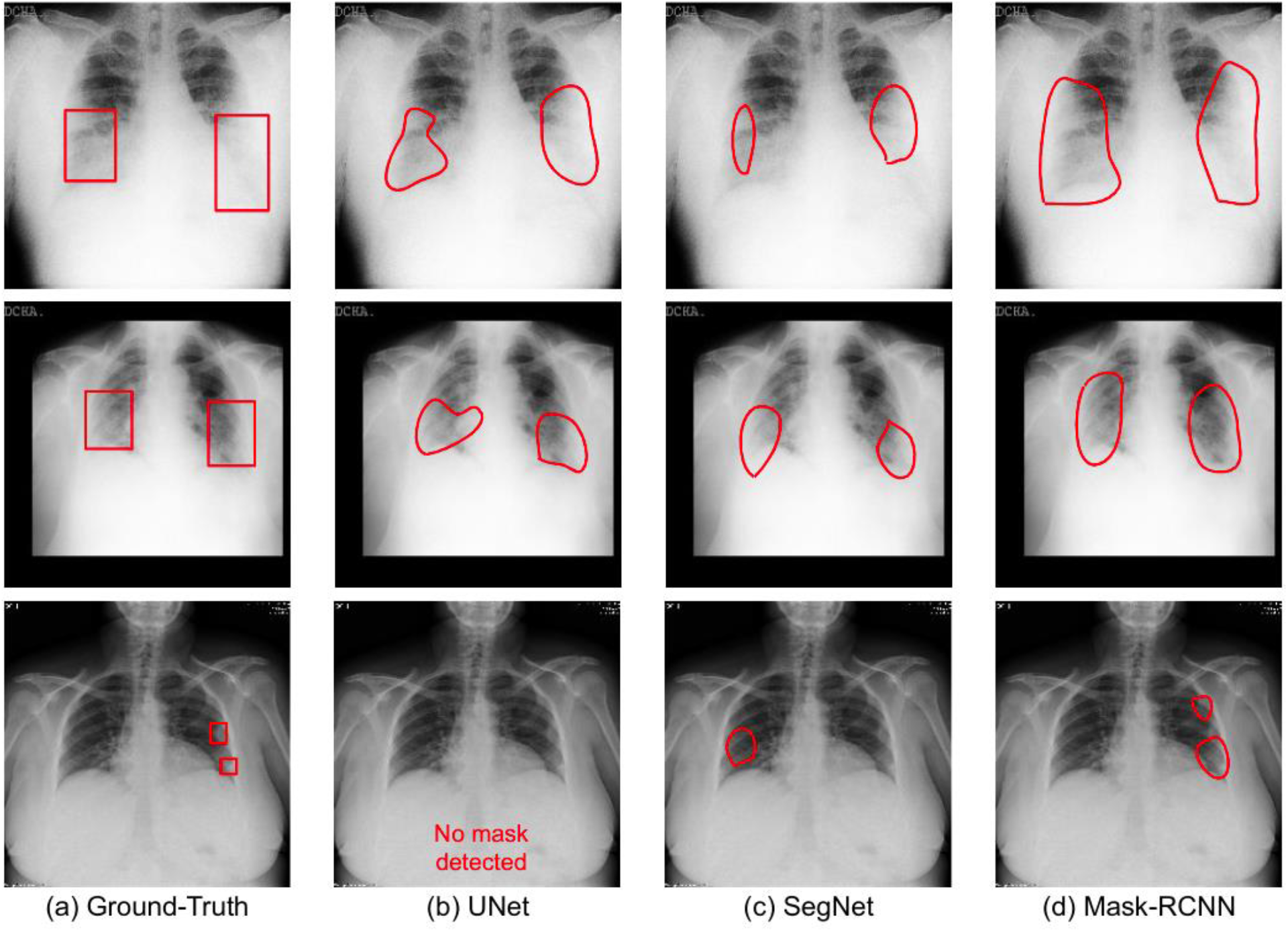
Samples of semantic disease segmentation for popular deep learning algorithms.

## Discussion

With the magnitude of Coronavirus pandemic, various deep learning algorithms aim to find the abnormalities in the lungs using CXRs, specific to COVID-19 pneumonia, and distinguish them from other etiologies of pneumonia. However, there are very few instances of public databases and to the best of our knowledge, there is no database with annotation of abnormalities on the chest x-rays of COVID-19 affected patients. In this work, we introduce the COVID Abnormality Annotation for X-Rays (CAAXR) database, built upon the BIMCV-COVID19+ database. Our major contribution to the database is the annotations of the abnormalities in frontal chest x-rays in the form of bounding boxes. Further, we also define four different sets of protocols for robust evaluation of the semantic segmentation and classification algorithms.

Finally, we benchmark the defined protocols and report the results using popular deep learning algorithms as a part of this study.

## Data Availability

There are two annotation files in this release. Filename - Study-Level_Annotations.csv, contains two types of study level annotations of these X-rays (Normal/Abnormal status & Image Quality). Filename - BoundingBox.csv, conatins pixel level image annotations for 10 Pathologies - Atelectasis, Cardiomegaly, Consolidation/Ground Glass opacity, Edema, Nodule, Pleural Effusion, Pleural Other, Pneumothorax. The corresponding X-rays were released by the Medical Imaging Data Bank of the Valencia region (BIMCV). They can be downloaded at the following link - https://bimcv.cipf.es/bimcv-projects/bimcv-covid19.

http://covbase4all.igib.res.in/

https://osf.io/b35xu/

## Key Points

- Provide bounding box abnormality annotation for COVID-19 positive chest x-rays.
- Propose semantic segmentation and classification protocols for a unified evaluation framework.
- Perform benchmarking for existing deep learning models on defined protocols.

## Abbreviations

(COVID-19): Coronavirus Disease 2019
(RT-PCR): real time polymerase chain reaction
(AI): artificial intelligence
(ROC): receiver operating characteristic
(AUC): area under the ROC curve
(CNN): convolutional neural network
(CXR): chest x-ray
(ML): machine learning

## Acknowledgements

This work is made possible by the tireless efforts of several organizations and team members, all of whom are mentioned on the CovBase Website. This specific annotation project has been made possible thanks to funding received from the Indian Institute of Technology, Jodhpur, INDIA.

**Data Availability Statement**

The data associated with this work is available here :

<u>Radiologists’ Annotations on COVID-19+ X-rays https://osf.io/b35xu/ via @OSFramework</u>

**Description**

Annotations by CARING’s (Centre for Advanced research in Imaging, Neuroscience and Genomics) expert radiologists on COVID-19+ X-rays as a part of the project titled “A Novel Abnormality Annotation Database forCOVID-19 Affected Frontal Lung X-rays” The corresponding X rays were released by Medical Imaging Data Bank of the Valencia region (BIMCV).

There are two annotation files in this release.

**Filename** - Study-Level_Annotations.csv, contains two types of study level annotations of these X rays (Normal/Abnormal status & Image Quality).

**Filename** - BoundingBox.csv, contains pixel level image annotations for 10 Pathologies - Atelectasis, Cardiomegaly, Consolidation/Ground Glass opacity, Edema, Nodule, Pleural Effusion, Pleural Other, Pneumothorax.

The corresponding X-rays were released by Medical Imaging Data Bank of the Valencia region (BIMCV). They can be downloaded at the following link - https://bimcv.cipf.es/bimcv-projects/bimcv/covid19.

